# Reliable estimation of SARS-CoV-2 anti-spike protein IgG titers from single dilution optical density values in serologic surveys

**DOI:** 10.1101/2022.02.13.22270904

**Authors:** Emilia M. M. Andrade Belitardo, Nivison Nery, Juan P. Aguilar Ticona, Moyra Machado Portilho, Guilherme S. Ribeiro, Mitermayer G. Reis, Federico Costa, Derek A. T. Cummings, Albert I. Ko, Mariam O. Fofana

**Affiliations:** Instituto Gonçalo Moniz, Fundação Oswaldo Cruz, Salvador BA, Brazil; Faculdade de Medicina, Universidade Federal da Bahia, Salvador BA, Brazil; Instituto de Saúde Coletiva, Universidade Federal da Bahia, Salvador BA, Brazil; Department of Biology, University of Florida, Gainesville FL, USA; Emerging Pathogens Institute, University of Florida, Gainesville FL, USA; Department of Epidemiology of Microbial Diseases, Yale School of Public Health, New Haven CT, USA

**Keywords:** SARS-CoV-2, serology, antibody, optical density, titers

## Abstract

**Background:** As the COVID-19 pandemic evolves, there is a need for reliable and scalable seroepidemiology methods to estimate incidence, monitor the dynamics of population-level immunity, and guide mitigation and immunization policies. Our aim was to evaluate the reliability of normalized ELISA optical density (nOD) at a single dilution as a predictor of SARS-CoV-2 immunoglobulin titers derived from serial dilutions.

**Methods:** We conducted serial serological surveys of a community-based cohort from the city of Salvador, Brazil after two sequential COVID-19 epidemic waves. Anti-SARS-CoV-2 spike protein immunoglobulin G (anti-S IgG) ELISA (Euroimmun AG) was performed with serial 3-fold dilutions of sera from 54 of the 1101 cohort participants. We estimated interpolated ELISA titers, used parametric models to fit the relationship between nOD at a single 1:100 dilution and interpolated titers, and assessed the correlation between changes in nOD and changes in titers.

**Results:** The relationship between nOD at a single 1:100 dilution and interpolated titers fit a log-log curve, with a residual standard error of 0.304. We derived a conversion table of nOD to interpolated titer values. Additionally, there was a high correlation between changes in nOD and changes in interpolated titers between paired serial samples (r = 0.836, ρ = 0.873). Changes in nOD reliably predicted increases and decreases in titers, with 98.1% agreement (κ = 95.9%).

**Conclusion:** Single nOD measurements can reliably estimate SARS-CoV-2 antibody titers, significantly reducing time, labor, and resource needs when conducting large-scale serological surveys to ascertain population-level changes in exposure and immunity.

**Highlights:**

- Optical density at a single dilution reliably estimates SARS-CoV-2 antibody titers
- Serial optical density measurements accurately identify changes in serostatus
- Using single optical density values can significantly reduce resource use in serosurveys

## Introduction

Previous studies have shown that the immune response to SARS-CoV-2 infection results in the development of multiple immunoglobulin classes (IgM, IgA and IgG) as early as the first week after the onset of symptoms ^1,2^. Serological assays are essential for epidemiological surveillance and to further the scientific understanding of SARS-CoV-2 immunity by monitoring the dynamics of population-level immunity as infections, vaccination and waning occur, and the resulting impact on transmission ^3–6^.

Whereas the qualitative presence or absence of antibodies provides meaningful information in non-immune individuals, in populations that have been highly exposed to infection and vaccination, ascertaining new infections requires assessing quantitative changes in antibody levels. The determination of binding antibody titers is typically very labor- and resource-intensive, as it requires measuring the presence of antibodies above a given threshold at multiple serial dilutions. Reducing the time and effort necessary for quantitation of antibody levels can help to expedite studies of immune response among individuals with exposure to SARS-CoV-2 vaccination or infection. Simpler and less costly methods of quantitation would be particularly valuable in resource-limited settings where laboratory capacity, staff, materials and reagents are scarce.

We therefore sought to assess whether the normalized ELISA normalized optical density (nOD) values at a single dilution could accurately estimate titers derived from serial dilutions.

Additionally, we evaluated the correlation between serial changes in nOD values and changes in titers.

## Materials and Methods

### Study site and population

This study was conducted with an open cohort of residents in the Pau da Lima community, which is located in Salvador, Brazil and for which long-term follow-up was conducted to study emerging infections ^7–10^. Household-based serological surveys have been conducted regularly at this site for several years. Individuals who sleep 3 or more nights per week within the defined study area, are aged 2 years or older, and who provide consent (parental consent for minors) were eligible to participate. Serological samples were collected from November 18 2020 to February 26 2021, after the first COVID-19 epidemic wave, and from July 14 2021 to October 31 2021, after the second wave, to evaluate seroprevalence and longitudinal trends in antibody response. A total of 1,101 individuals had paired longitudinal samples from both periods. We selected a sample of 54 individuals, aiming to achieve representation of a broad range of nOD and titer values to fully characterize the relationship between these measurements (Figure 1). This sample included 48 individuals who were seropositive during the first study period, and 18 individuals who were vaccinated prior to the second study period. Additionally, 195 banked samples collected from cohort participants between September 9 and November 11 2019, prior to the emergence of COVID-19 in Brazil, served as negative controls.

**Figure 1:**
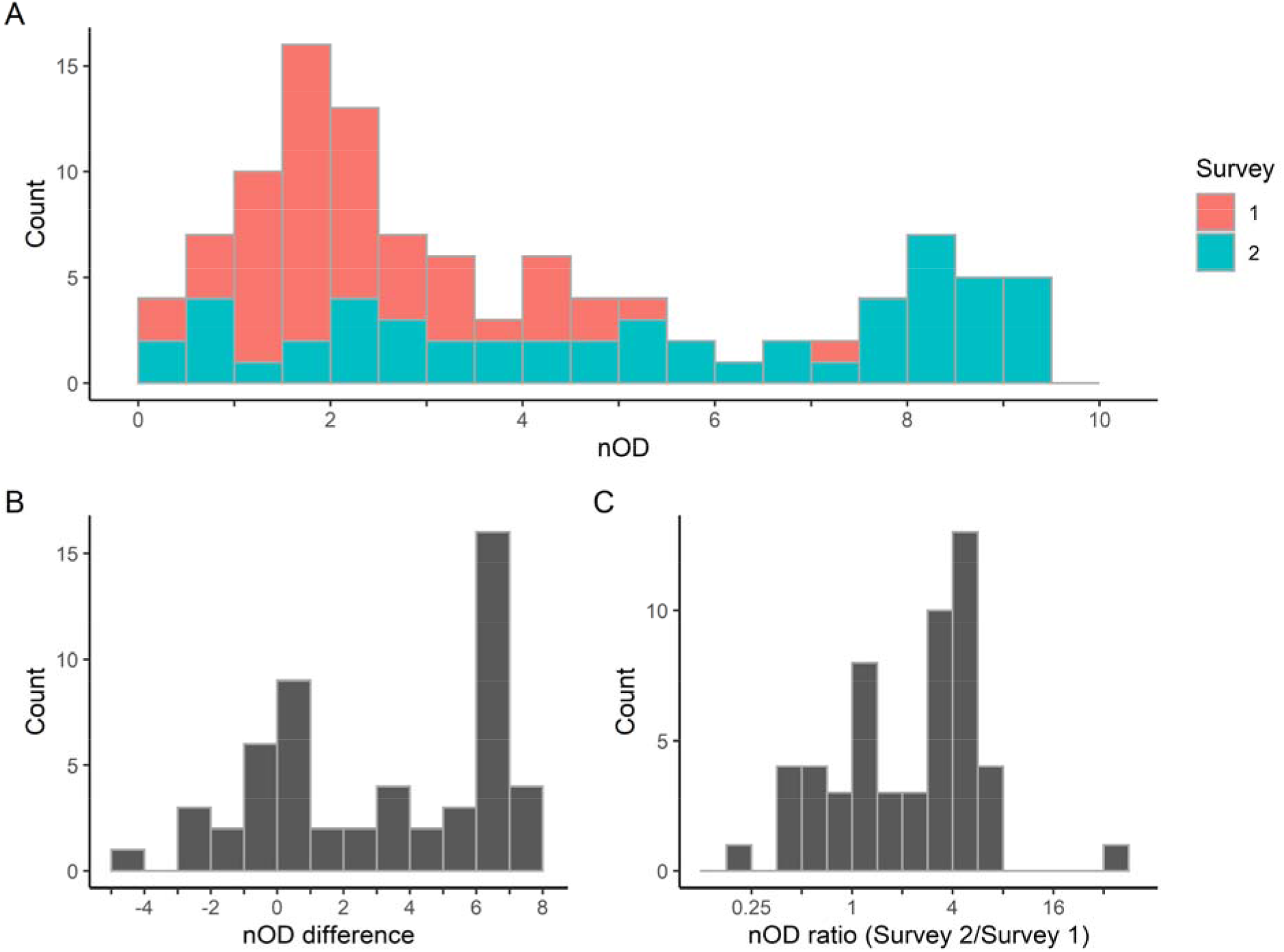
Distribution of samples selected for titer measurements. A: nOD values of samples collected during survey 1 (pink) and survey 2 (green). B: difference in nOD values between survey 1 and survey 2. C: ratio of nOD values (survey 2: survey 1).

### Ethical and confidentiality considerations

Study participants were informed about the project, the risks and the absence of immediate individual benefits. Participation in the study was voluntary and could be interrupted at any time. All adult participants signed an informed consent form in the presence of witnesses prior to enrollment, in accordance with Resolution no. 466/2012 of the Brazil Ministry of Health. Parental consent was obtained for minors. This project was approved by the Human Research Ethics Committee of the Instituto Gonçalo Moniz, Fundação Oswaldo Cruz (FIOCRUZ), the National Research Ethics Council (CONEP) and the Yale University Research Ethics Committee.

### Collection, transport and storage of samples

Trained members of the study team performed venipuncture. Blood samples were collected in a dry tube and transported to FIOCRUZ in refrigerated transport boxes. After centrifugation at 3000 RPM (1,811 RCF) at 4°C for 15 minutes, the samples were aliquoted and stored at -20°C.

### SARS-CoV-2 IgG ELISA and titers

Serological assays for detection of IgG against the SARS-CoV-2 spike protein were performed using commercial ELISA kits (Euroimmun AG, Lübeck, Germany) and plates pre-coated with the S1 protein. Samples were diluted 1:101 in buffer and processed according to the manufacturer’s instructions. Briefly, 100µl of each sample, calibrator, and positive and negative controls were added to the plate and incubated for 1 hour at 37°C. After three wash steps with wash buffer, 100µl of HRP-labeled secondary anti-human IgG was added for 30 minutes at 37°C. The plates were washed three more times with wash buffer and 100µl of substrate solution (TMB/H_2_O_2_) was added for 30 minutes at room temperature, with shielding from light. The reaction was stopped with the addition of 100µl of 0.5M sulfuric acid and the absorbance was measured at a wavelength of 450nm using an automated plate reader (Tecan Austria GmbH, GrLJdig, Austria). Normalized optical density (OD) values were calculated as the ratio of the OD of each test sample to that of the calibrator.

Titers were obtained by qualitative assessment of antibody binding at five serial 3-fold dilutions (1:100, 1:300, 1:900, 1:2700, 1:8100), based on previously published protocols ^11^. Interpolated titers were computed using the software GraphPad Prism (Version 5.01, GraphPad software, San Diego, USA). These values correspond to the estimated titer at which the presence of antibody is no longer detected, interpolated from the highest dilution with positive antibody detection and the next serial dilution. Unlike endpoint titers, which are interval-censored, interpolated titers are on a continuous scale, and result in less biased estimates ^12^. All assays were conducted by a single operator, on 35 distinct days.

### Threshold for antibody detection

For the Euroimmun anti-S IgG ELISA, the manufacturer recommends that samples with normalized OD values <0.8 are considered negative, those with values >=0.8 and <1.1 considered borderline, and those with values >=1.1 considered positive. We defined samples with values ≥ 0.8 as positive. In order to evaluate the appropriateness of these cutoffs, which were derived from evaluation of COVID-19 patients, to the context of a seroprevalence survey in our study population, we performed assays using pre-pandemic serum samples as negative controls. We then applied a widely accepted method to establish cutoffs in the absence of known positive standards, using the upper prediction limit from negative samples ^13^.

### Statistical analysis

We compared interpolated titers to the normalized OD obtained at the 1:100 dilution. The relationship between these values fit a sigmoidal function, consistent with previous observations ^14^. We fit a 5-parameter log-log curve to the data using the R packages “aomisc” and “drc” ^15,16^.

We assessed both the Pearson and Spearman (rank) correlation coefficients for the change in normalized OD and change in interpolated titers. Given that a 4-fold change in titers is a common criterion to identify recent antigen exposure such as would occur with incident infection ^17,18^, we computed the area under the ROC curve for the change in nOD values that would correspond to 4-fold increases or decreases in interpolated titers. All analyses were conducted using the software R, version 4.1.1 ^19^.

## Results

### Estimation of interpolated titers using normalized OD

In our primary analysis we estimated interpolated titers using a normalized OD cutoff of 0.8. The relationship between interpolated titers and normalized OD at a single 1:101 dilution exhibited a clear sigmoidal curve pattern. We fitted the data to a 5-parameter log-log curve, with the following form: 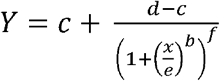,where *c* and *d* are the lower and upper asymptotes, respectively, and *f* is an asymmetry parameter. Parameters *b* and *e* characterize the Hill’s slope and inflection point. The residual standard error (RSE) of the fitted values was 0.304. Details of the model parameters are shown in Table 2.

**Table 2:**
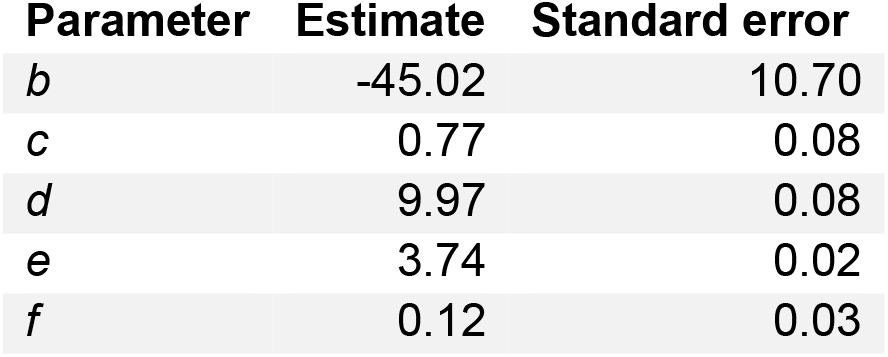
Curve fit parameters.

### Prediction of changes in serostatus and antibody titers

We compared the observed changes in interpolated titers to the changes in normalized OD at a single 1:100 dilution (Figure 2). The estimated interpolated titers corresponding to a wide range of nOD values are shown in Table 3.

**Figure 2:**
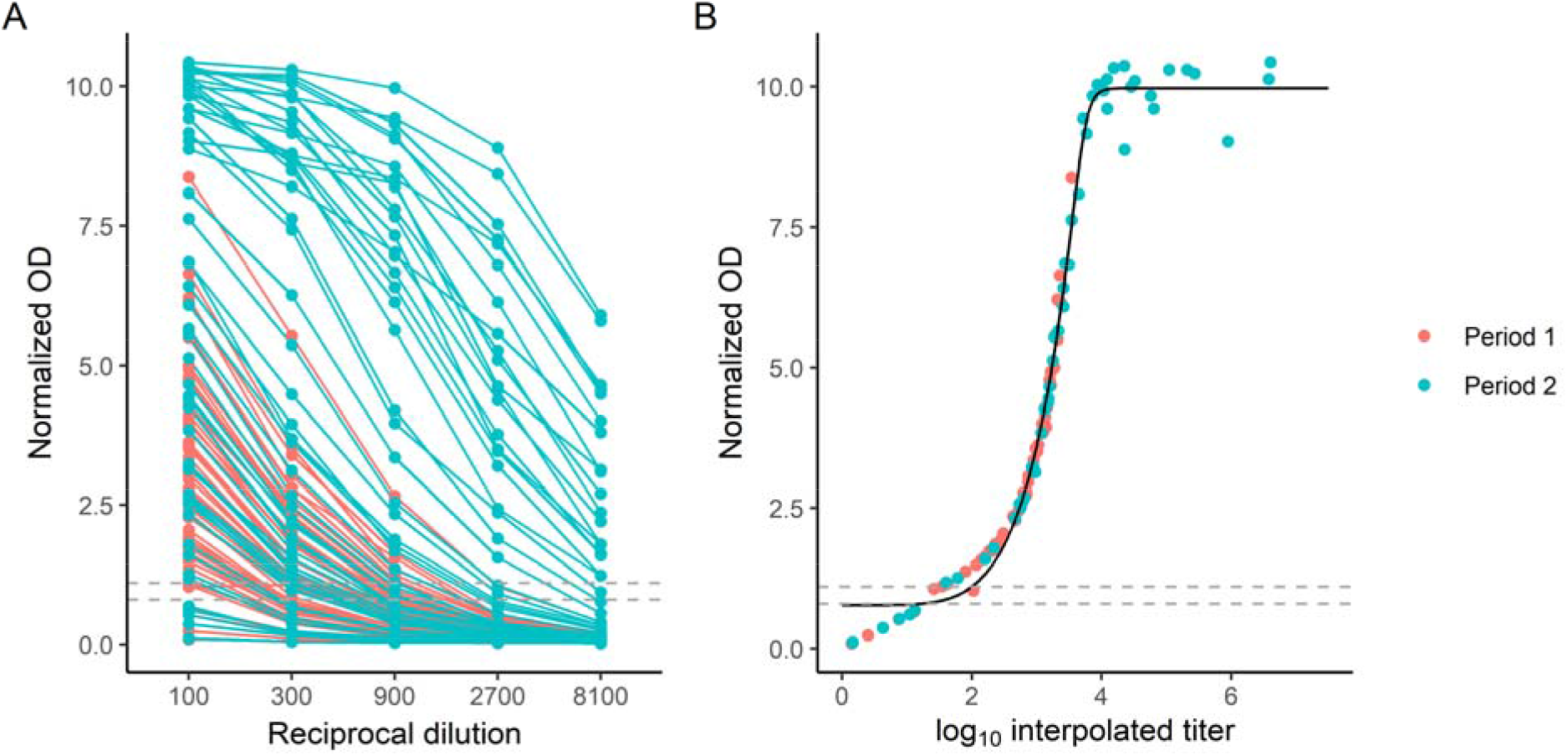
Anti-S IgG measurements. A: Summary of normalized OD values obtained at each serial dilution. Each curve represents a single sample collected during period 1 (pink) or period 2 (green). The dashed gray lines indicate the manufacturer-suggested cutoffs of 0.8 and 1.1. B: Comparison of the interpolated titer as estimated from serial dilutions to the normalized OD measured at a single dilution of 1:100 revealed a sigmoidal relationship. Dashed gray lines represent the manufacturer-suggested cutoffs of 0.8 and 1.1. The solid black line shows fitted values based on a 5-parameter log-log curve.

**Table 3:** ELISA nOD to titer conversion.

| **nOD** | **Estimated titer (95%CI)** | **nOD** | **Estimated titer (95% CI)** |
| --- | --- | --- | --- |
| 0.8 | 0.93 (0.82-1.03) | 5.4 | 3.29 (3.27-3.31) |
| 0.8 | 1.24 (1.13-1.34) | 5.5 | 3.3 (3.28-3.32) |
| 0.9 | 1.67 (1.57-1.77) | 5.6 | 3.31 (3.29-3.33) |
| 1 | 1.86 (1.77-1.96) | 5.7 | 3.33 (3.31-3.35) |
| 1.1 | 2 (1.91-2.09) | 5.8 | 3.34 (3.32-3.36) |
| 1.2 | 2.1 (2.02-2.19) | 5.9 | 3.35 (3.33-3.37) |
| 1.3 | 2.19 (2.1-2.27) | 6 | 3.36 (3.35-3.38) |
| 1.4 | 2.26 (2.18-2.34) | 6.1 | 3.38 (3.36-3.4) |
| 1.5 | 2.32 (2.25-2.4) | 6.2 | 3.39 (3.37-3.41) |
| 1.6 | 2.38 (2.31-2.45) | 6.3 | 3.4 (3.38-3.42) |
| 1.7 | 2.43 (2.36-2.5) | 6.4 | 3.41 (3.39-3.43) |
| 1.8 | 2.48 (2.41-2.54) | 6.5 | 3.42 (3.4-3.44) |
| 1.9 | 2.52 (2.46-2.58) | 6.6 | 3.44 (3.41-3.46) |
| 2 | 2.56 (2.5-2.62) | 6.7 | 3.45 (3.43-3.47) |
| 2.1 | 2.6 (2.54-2.66) | 6.8 | 3.46 (3.44-3.48) |
| 2.2 | 2.64 (2.58-2.69) | 6.9 | 3.47 (3.45-3.49) |
| 2.3 | 2.67 (2.62-2.72) | 7 | 3.48 (3.46-3.5) |
| 2.4 | 2.7 (2.65-2.75) | 7.1 | 3.49 (3.47-3.51) |
| 2.5 | 2.73 (2.68-2.78) | 7.2 | 3.5 (3.48-3.52) |
| 2.6 | 2.76 (2.71-2.81) | 7.3 | 3.51 (3.49-3.53) |
| 2.7 | 2.79 (2.74-2.84) | 7.4 | 3.52 (3.5-3.55) |
| 2.8 | 2.82 (2.77-2.86) | 7.5 | 3.53 (3.51-3.56) |
| 2.9 | 2.84 (2.8-2.88) | 7.6 | 3.54 (3.52-3.57) |
| 3 | 2.87 (2.82-2.91) | 7.7 | 3.55 (3.53-3.58) |
| 3.1 | 2.89 (2.85-2.93) | 7.8 | 3.57 (3.54-3.59) |
| 3.2 | 2.91 (2.87-2.95) | 7.9 | 3.58 (3.55-3.6) |
| 3.3 | 2.93 (2.9-2.97) | 8 | 3.59 (3.56-3.61) |
| 3.4 | 2.96 (2.92-2.99) | 8.1 | 3.6 (3.57-3.62) |
| 3.5 | 2.98 (2.94-3.01) | 8.2 | 3.61 (3.58-3.64) |
| 3.6 | 3 (2.96-3.03) | 8.3 | 3.62 (3.59-3.65) |
| 3.7 | 3.02 (2.98-3.05) | 8.4 | 3.63 (3.6-3.66) |
| 3.8 | 3.04 (3-3.07) | 8.5 | 3.64 (3.61-3.67) |
| 3.9 | 3.05 (3.02-3.09) | 8.6 | 3.65 (3.62-3.69) |
| 4 | 3.07 (3.04-3.1) | 8.7 | 3.67 (3.63-3.7) |
| 4.1 | 3.09 (3.06-3.12) | 8.8 | 3.68 (3.64-3.71) |
| 4.2 | 3.11 (3.08-3.14) | 8.9 | 3.69 (3.65-3.73) |
| 4.3 | 3.12 (3.1-3.15) | 9 | 3.7 (3.66-3.75) |
| 4.4 | 3.14 (3.12-3.17) | 9.1 | 3.72 (3.67-3.76) |
| 4.5 | 3.16 (3.13-3.18) | 9.2 | 3.73 (3.68-3.78) |
| 4.6 | 3.17 (3.15-3.2) | 9.3 | 3.75 (3.7-3.8) |
| 4.7 | 3.19 (3.16-3.21) | 9.4 | 3.77 (3.71-3.83) |
| 4.8 | 3.2 (3.18-3.23) | 9.5 | 3.79 (3.72-3.86) |
| 4.9 | 3.22 (3.2-3.24) | 9.6 | 3.82 (3.74-3.89) |
| 5 | 3.23 (3.21-3.25) | 9.7 | 3.85 (3.76-3.93) |
| 5.1 | 3.25 (3.23-3.27) | 9.8 | 3.89 (3.78-4) |
| 5.2 | 3.26 (3.24-3.28) | 9.9 | 3.98 (3.83-4.12) |
| 5.3 | 3.27 (3.25-3.3) | 10 | 4.17 (3.93-4.42) |

Overall, there was a good correlation between the difference in log interpolated titer and the difference in normalized OD between paired samples from study periods 1 and 2 (Pearson correlation r^2^ = 0.78, Spearman rank correlation ρ = 0.868; Figure 3).

**Figure 3:**
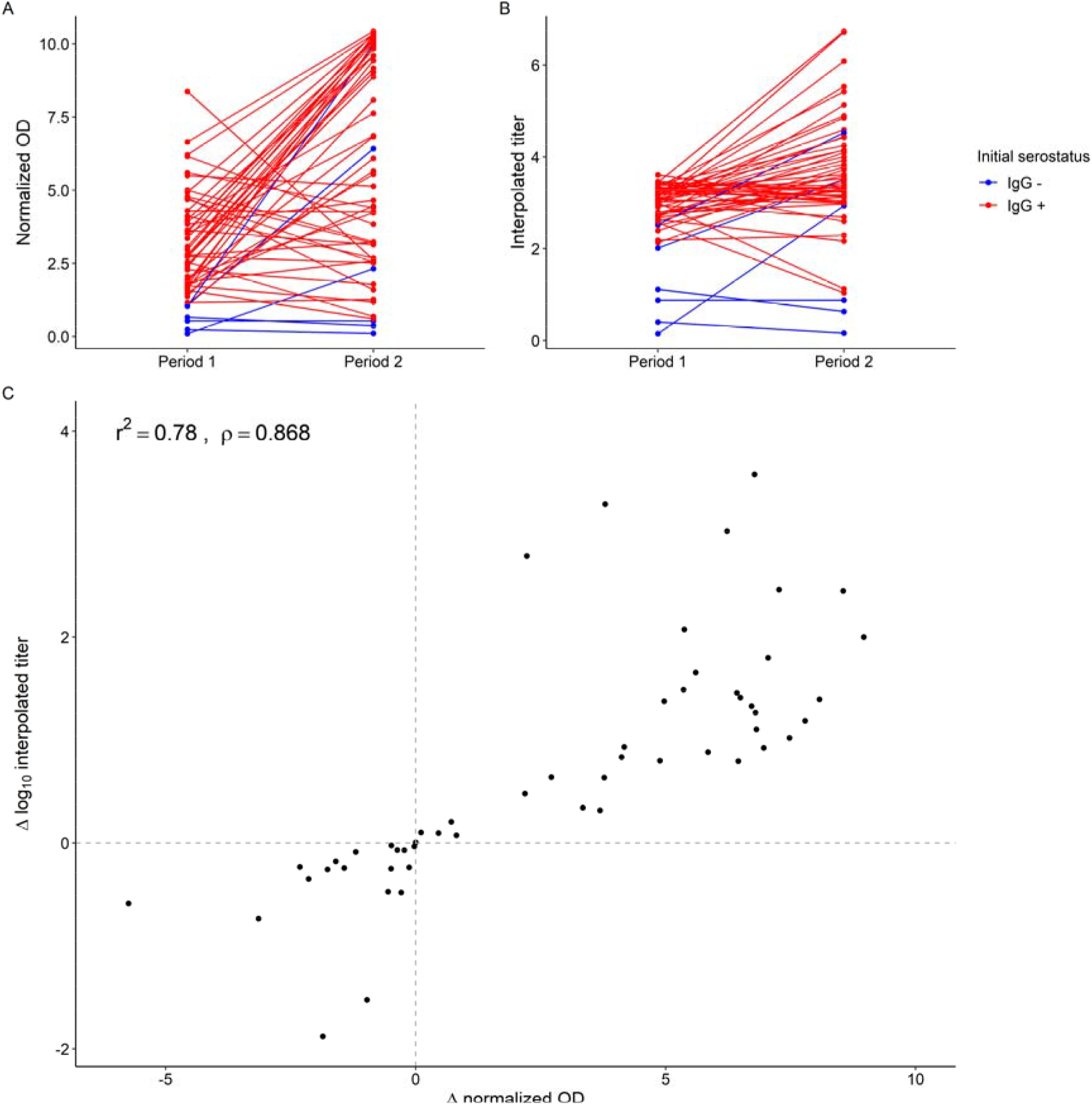
Change in normalized OD and titers. Spaghetti plots of individual change in normalized OD (A) and interpolated titer (B) between period 1 and period 2. Panel C illustrates the concordance in the direction of change (increase vs. increase) and the correlation between the difference in log interpolated titer and the difference in normalized OD at a single 1:100 dilution. Each data point represents the difference between study periods 1 and 2 for the same individual. Dashed gray lines indicate no change in OD or titer.

There was 98.1% concordance (κ = 95.9%) for the detection of an increase or decrease in interpolated titer. The area under the receiver operating characteristic curve (AUC) values for the detection of a four-fold increase or decrease in interpolated titer was 0.994 (95% confidence interval [CI] 98.4-100.0%). A 1.48-fold change in nOD predicted a 4-fold rise in interpolated titer with 100% sensitivity and 92% specificity. A 2-fold change in nOD was more specific (100%) but less sensitive (93.1%). We repeated our analyses using a cutoff of 1.1 for presence of anti-S1 antibodies and did not observe any significant changes to our findings (Figure 4).

**Figure 4:**
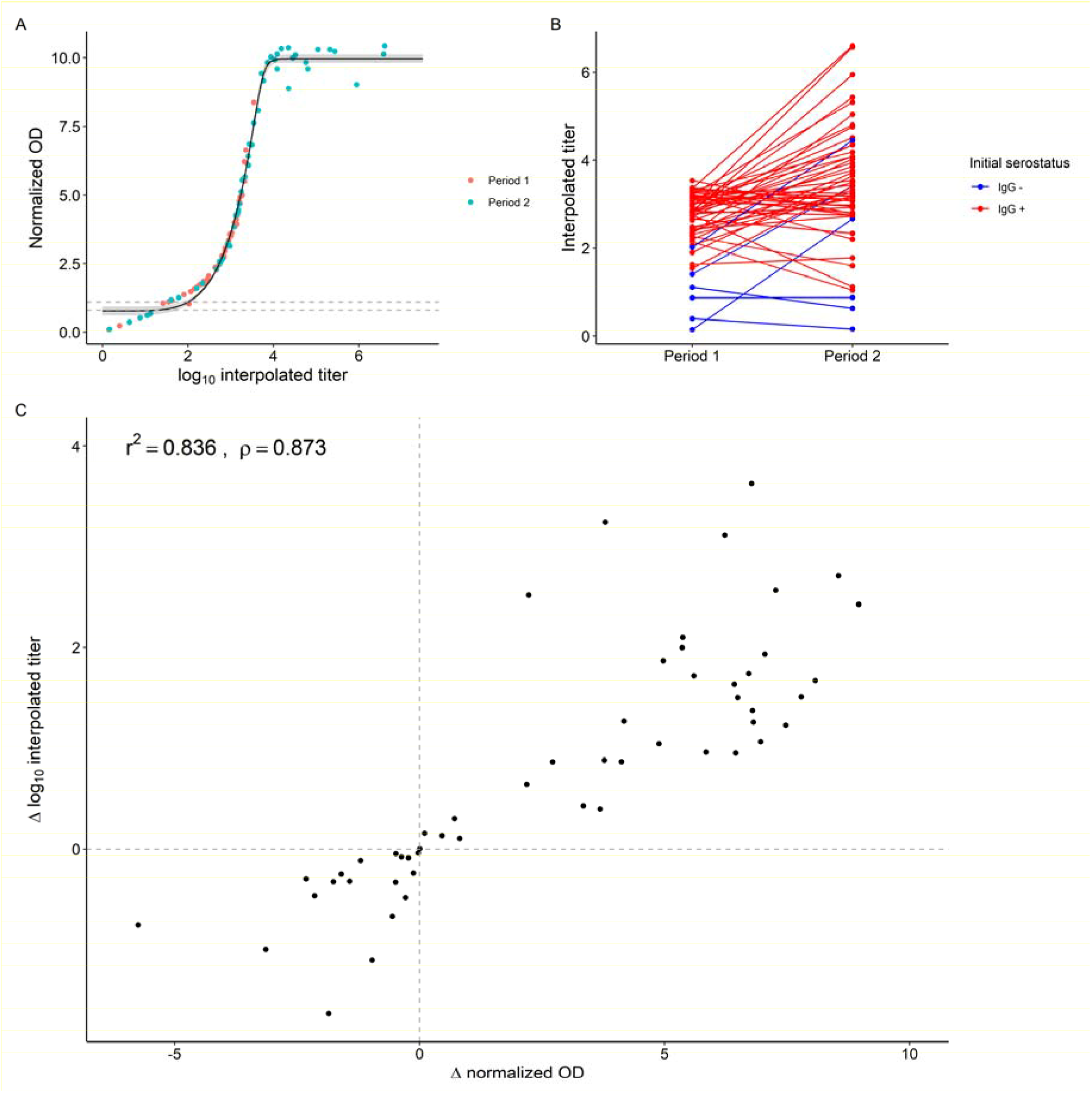
Sensitivity analyses. We repeated our primary analyses using an OD cutoff for the presence of anti-S antibody of 1.1 (vs. 0.8). The relationship and parametric fit of normalized OD values to interpolated titers (A), as well as the correlation between the difference in normalized OD and difference in interpolated titer (C) remained similar (r^2^ = 0.836, ρ = 0.873).

## Conclusions

We found a predictable correlation between ELISA nOD values at a single 1:100 dilution and interpolated titers derived from serial dilutions. We were able to fit the relationship between normalized OD and interpolated titers to a parametric log-log curve, such that OD values could be used to estimate corresponding interpolated titers. Our results demonstrate that a single ELISA optical density measurement using a widely available commercial assay can reliably estimate SARS-CoV-2 IgG titers for population-level serological surveys. Moreover, we found a high correlation between changes in normalized OD and changes in interpolated titers between paired serial samples from the same individuals.

One limitation is that we did not assess the correlation between OD values and virus neutralization activity. Nevertheless, prior studies have demonstrated that binding antibody levels correlate well with neutralizing activity ^20^. Although assessment of neutralizing activity is necessary in certain contexts, binding antibody levels are more practical and scalable for applications such as population serological surveys, or assessment of immune response to vaccination. Our findings raise the possibility that future studies of antibody waning and response to booster vaccine doses could rely on a single ELISA OD measurement of paired samples rather than requiring serial titration of each sample, thus greatly reducing the necessary effort and expense. Moreover, our study population included individuals with a broad range of binding antibody levels, allowing us to characterize the full range of optical density values up to the upper limit of quantitation.

Another limitation of our study is that these assays were performed in a single lab and by a single operator. There may be additional variability across operators and laboratories. Reassuringly, despite using samples that were collected 6 months apart and processed over several weeks, there was no significant systematic variation. Future studies should assess the validity of our estimation tool in other settings, to validate its applicability.

There is a continuing need to expand access to SARS-CoV-2 research capability in low- and middle-income countries. Serological surveys are especially important in settings where there may be a lower proportion of symptomatic infections (i.e., younger populations), as they can identify infections that would otherwise go undetected. With the continuing rollout of vaccination and boosters, serological surveys will play an important role in research investigating the dynamics of population-level immunity and the resulting impact on transmission. Serial measurements, or combined measurement of IgG and other immunoglobulins, could serve to reconstruct the infection history and immunity dynamics of populations to guide policies on mitigation measures and vaccination (e.g., number of doses, timing, priority populations). Our findings demonstrate that such studies can be conducted with a single ELISA nOD measurement, thus reducing the effort, time, and cost involved. We expect that these gains will be particularly valuable in resource-limited settings where laboratory capacity is strained.

## Data Availability

All data produced in the present study are available upon reasonable request to the authors

